# Brain Structure and Substance Use: Disentangling Risk, Exposure, and Drug-Specific Effects

**DOI:** 10.64898/2026.03.20.26348897

**Authors:** Daniella A Fernandez, David AA Baranger

## Abstract

**Importance:** Polysubstance use is common, but substance use associations with neuroimaging measures have largely been investigated within individual drug types. Whether effects are substance-specific or -general, and how predispositional risk and exposure contribute, remains unclear.

**Objective:** Identify shared and unique associations between substance use and brain structure, and characterize the contributions of predispositional risk and environmental exposure, in a large sample of young adults in the US.

**Design:** This cross-sectional family-based study used data from the Human Connectome Project (2017 release, collected from 2012-2015).

**Setting:** Data were collected at Washington University in St. Louis, MO, USA.

**Participants:** Twins, non-twin siblings, and singletons with magnetic resonance imaging (MRI) and substance use self-report were included in the analysis. Data were analyzed in 2025.

**Exposure:** History of substance use was assessed using the Semi-Structured Assessment for the Genetics of Alcoholism. Variables included lifetime use, heavy or past-year hazardous use, and age of use onset for alcohol, marijuana, tobacco, and illicit drugs. Additionally, alcohol and marijuana dependence were assessed.

**Main Outcomes and Measures:** Linear mixed-effect models examined associations between substance use and brain structure, with an initial focus on past-year hazardous alcohol use, as 95% of the sample endorsed lifetime alcohol use. Analyses then tested associations with other substance use variables, and whether effects were shared or substance-specific. Between-family, within-family, and genetic variance component analyses tested risk and exposure effects.

**Results:** 1,113 participants (N = 445 families; ages 22 - 37; *M=*28.8, *SD*=3.7) had no missing data for the primary analyses. Hazardous alcohol use was negatively associated with global brain thickness (*β* = −0.12, *p* < 0.001), which explained all other regional and global associations. Of the drugs with a shared-effect on global brain thickness, only lifetime marijuana use explained unique variance over alcohol (*β* = −0.08, *p* = 0.013). Within-family analyses found evidence for unique putative exposure effects of both alcohol (*β* = −0.11, *p* < 0.001) and marijuana use (*β* = −0.07, *p* = 0.002) on global thickness. Marijuana use further showed a predispositional effect, both in between-family comparisons (*β* = −0.11, *p* = 0.007) and genetic variance component analyses (*ρG* = −0.2*, p* = 0.004), which were not explained by alcohol use.

**Conclusions and Relevance:** Brain structural associations with substance use reflect substance-general and -specific effects, as well as a combination of predispositional and exposure effects. Findings suggest that the negative consequences of polysubstance use may reflect the additive effects of multiple unique exposures.

## Introduction

Approximately 20% of Americans engage in polysubstance use, or the use of drugs pertaining to more than one drug class ^1^. These individuals experience higher rates of suicidality, incarceration, and mental health comorbidities compared to monosubstance users ^2–6^. Additionally, 58% of lethal drug overdoses in the United States involve polysubstance use ^7^.

Despite the prevalence of polysubstance users and the intensity of their condition, this population is largely underrepresented in the psychiatric neuroimaging literature. Indeed, substance use associations with neuroimaging measures have largely been investigated within individual drug types. These robust and well-replicated studies often report differences in brain structure that overlap across alcohol, marijuana, tobacco, and illicit drug use, suggesting shared correlates or predispositional mechanisms ^8–11^. Emerging work has examined a combination of these substance types in tandem and consistently identified shared-drug effects on brain volume and thickness ^9,12–16^.

Beyond the effects of shared use, less is known about unique-drug effects, where individual associations are not explained by broader polysubstance use. This growing literature has emphasized comparisons of alcohol use to other drug classes, likely as alcohol is the most widely used substance ^2^. Unique-drug effects of alcohol use have been frequently observed over marijuana, tobacco, and illicit drug use across global and regional brain structures ^12,17–20^. Additional data suggest that brain structure associations with marijuana and tobacco are not substance-specific ^15,16,19,21,22^. However, prior investigations of shared- and unique-effects have mostly examined only two or three drug classes, and less is known about how different dimensions of use compare (e.g., age of onset, recent use, hazardous use, dependence).

Well-established neurobiological models of addiction posit the influence of both substance exposure and risk for use on the brain. Neurodevelopmental and genetic risk models emphasize how pre-existing risk, including genetics and shared environment, influence the development and functioning of neural circuitry underlying substance use propensity, initiation, and other dimensions of use ^11,23,24^. Stage-based models then describe how, upon initiation and continued use, this neural circuitry undergoes further changes that promote stages of addiction: intoxication, negative reinforcement, and craving ^25,26^. Neuroimaging studies with family-based data, longitudinal designs, and genetic analyses have successfully built on these conceptual frameworks and identified predispositional risks and substance-exposure effects on brain structure ^24^. However, while both risk-based and stage-based neurobiological models emphasize mechanistic pathways that are substance-general, it remains unknown whether the predispositional and exposure-related associations of substance use with brain structure reflect drug-shared or -unique effects.

The Human Connectome Project (HCP) is a prominent family-based data set used in prior work to investigate associations of drug use and brain structure ^10,27–31^. However, the entire range of substance use data present in the sample has not been fully explored, particularly in concert with analyses that leverage the family design. Here we examine structural associations with multiple dimensions of alcohol, marijuana, tobacco, and illicit drug use to identify drug-shared and -specific effects. Further, we use genetic and environmental (dis)similarities between siblings to disentangle influences of exposure effects and predispositional risk in that association. These data further our understanding of factors that influence the relationship between substance use and the human brain.

## Methods

Data were drawn from the 1,200 Subjects release of the Human Connectome Project (HCP), a cross-sectional family-based study of individual differences in brain circuits and their relation to behavior and genetic background ^27,31–33^. 1,113 participants who had complete data for the primary variables and covariates were included in analyses. Participant age ranged from 22 to 37 years old (*M_age_ =* 28.86, *SD_age_* = 3.7), and 54% (*n* = 606) were female (**Table 1**). For further details (e.g., inclusion criteria, study procedures), see **eMethods**.

**Table 1.** Sample Demographics.

|  | *M (SD; Range)* |
| --- | --- |
| Age in Years | 28.8 (3.7; 22-37) |
|  | *n (%)* |
| Gender |  |
| Male | 507 (46) |
| Female | 606 (54) |
| Race |  |
| White | 830 (74.5) |
| Black/ African American | 168 (15.1) |
| Asian/Native Hawaiian/Other Pacific Islander | 65 (5.8) |
| American Indian/Alaskan Native | 2 (0.2) |
| More than one | 29 (2.6) |
| Unknown/Not reported | 19 (1.7) |
| Income |  |
| <\$10,000 | 80 (7.2) |
| \$10K–19,999 | 88 (8) |
| \$20K–29,999 | 138 (12.4) |
| \$30K–39,999 | 132 (11.9) |
| \$40K–49,999 | 115 (10.3) |
| \$50K–74,999 | 232 (20.8) |
| \$75K–99,999 | 149 (13.4) |
| ≥\$100,000 | 172 (15.5) |
| Not Reported | 7 (0.6) |
| Education (Years Completed) |  |
| <11 | 39 (3.5) |
| 12 | 156 (14) |
| 13 | 69 (6.2) |
| 14 | 137 (12.3) |
| 15 | 69 (6.2) |
| 16 | 467 (42) |
| 17+ | 174 (15.6) |
| Not Reported | 2 (0.2) |
| Sibling Status |  |
| Monozygotic Twin | 306 (27.5) |
| Dizygotic Twin | 173 (15.5) |
| Half | 27 (2.4) |
| Non-Twin Sibling | 543 (47.8) |
| Singleton | 64 (5.8) |

### Measures

#### Lifetime Drug Use and Dependency

Participants were administered the Semi-Structured Assessment for the Genetics of Alcoholism (SSAGA) ^34^, capturing information on their history of alcohol, marijuana, tobacco, and illicit drug (i.e., cocaine, stimulant, sedative, opiates, other) use. Binary lifetime use and age of use onset were recorded for all drug types. Heavy marijuana and tobacco users were defined as having used marijuana more than 1,000 times and smoking more than 100 cigarettes, respectively. Maximum illicit drug use was indexed as the longest period of almost every day use. Lifetime drug dependency, as outlined by the Diagnostic and Statistical Manual of Mental Disorders-4 (DSM-4), was assessed for marijuana and alcohol use.

#### Hazardous Alcohol Consumption

As in prior work ^28^, a measure of hazardous alcohol use, mAUDIT-C, was generated by summing responses to three items of the SSAGA that are highly similar to the three items of the AUDIT-C ^35^, a well-validated and widely used measure ^36^. For additional details on scoring transformations, see **eMethods** and **eTable 1**.

#### Brain Structure

High-resolution (0.7-mm isotropic voxels) anatomical images were acquired using a customized Siemens Skyra 3-T scanner with a 32-channel head coil ^31,33^. For details on imaging procedures and processing, see **eMethods**.

#### Covariates

Participants self-reported biological sex, age, socioeconomic status, and level of educational attainment. Sibling status (i.e., monozygotic twin, dizygotic twin, half) was identified via genetic testing and self-report. Additionally, intracranial volume, excluding the ventricles, was included in all analyses.

### Statistical Analysis

Linear mixed effects models tested the association of substance use variables with brain structure ^37,38^, including a random intercept for Family ID and fixed effect covariates for age, age^2^, sex, socioeconomic status, level of educational attainment, intracranial volume, and sibling status ^27,39^. Data were scaled, centered, and winsorized (+/− 3 SD) to obtain standardized estimates and reduce the influence of extreme values. Skewed variables (skew >1, e.g., times used illicit drugs, marijuana dependency) were log-transformed to reduce the influence of extreme values. FDR multiple-test correction ^40^ was used throughout.

Analyses first tested associations with the mAUDIT-C, as 95% of the sample endorsed lifetime alcohol use. Regions were selected based on a prior review that identified brain measures with consistent evidence for predispositional and/or causal associations with alcohol use ^24^. Analyses identified global brain thickness as the strongest association, which, when included as an additional covariate, accounted for all other associations (**Results**). *Post-hoc* analyses tested associations with all regions (**eMethods; eResults**). Shared-effects of drug use on brain thickness were examined by testing associations with hazardous alcohol and lifetime marijuana, tobacco, and illicit drug use. Other variables reflecting degrees of use (i.e., age of onset, dependence, heavy use) were tested with lifetime use as a covariate. Unique effects were then identified by incorporating all variables that were individually significant in a single model.

Predispositional and exposure effects of drug use and brain thickness were examined with between/within family analyses. Between-family effects reflect predispositional risk, as they capture shared risk factors that differ between families ^41^. Within-family effects are consistent with exposure risk, as they are sensitive to differences in environment and non-shared genetics that differ between siblings and twins^42,43^. Between- and within-family drug use variables were computed by calculating the family average of each substance use variable, and then individual deviations from the family average, respectively. To assess bidirectional associations of brain structure and substance use, predictor and response variables of the previous analyses were reversed (i.e., between- and within-family brain thickness fit to predict substance use variables). To probe the robustness of these effects, all between-and within-analyses were repeated in increasingly stringent, genetically similar subsamples (i.e., full-siblings only, twins only, and monozygotic twins only). A secondary test of predispositional effects was conducted using the SOLAR-Eclipse Genetics software (https://solar-eclipse-genetics.org/) ^44^, which leverages differences between monozygotic and dizygotic/non-twin siblings to compute heritability, non-shared environment, and variance component correlation estimates.

## Results

### Sample Characteristics

The sample consisted of 1,113 participants who were, on average, 28.8 years old (*SD* = 3.7). The participant pool was primarily female (*n* = 606, 54%) and White (*n* = 830, 74.6%) (**Table 1; eFigure 1**). 1,053 participants (95%) endorsed lifetime alcohol use, 564 participants (51%) endorsed lifetime marijuana use, 480 participants (43%) endorsed lifetime tobacco use, and 221 (20%) endorsed lifetime illicit drug use. 726 (65%) endorsed lifetime use of two or more substances and 134 (12%) endorsed lifetime hazardous use of two or more substances (**eFigure 2**). Of the participants who endorsed lifetime alcohol use, less than half were categorized as low hazardous alcohol use ^45^ (*n* = 479, 45%) (**Figure 1**). Further information on substance use endorsement, including patterns of hazardous use and age of use onset, is shown in **eTable 2**, **eFigure 2, and eFigure 3**.

**Figure 1.**
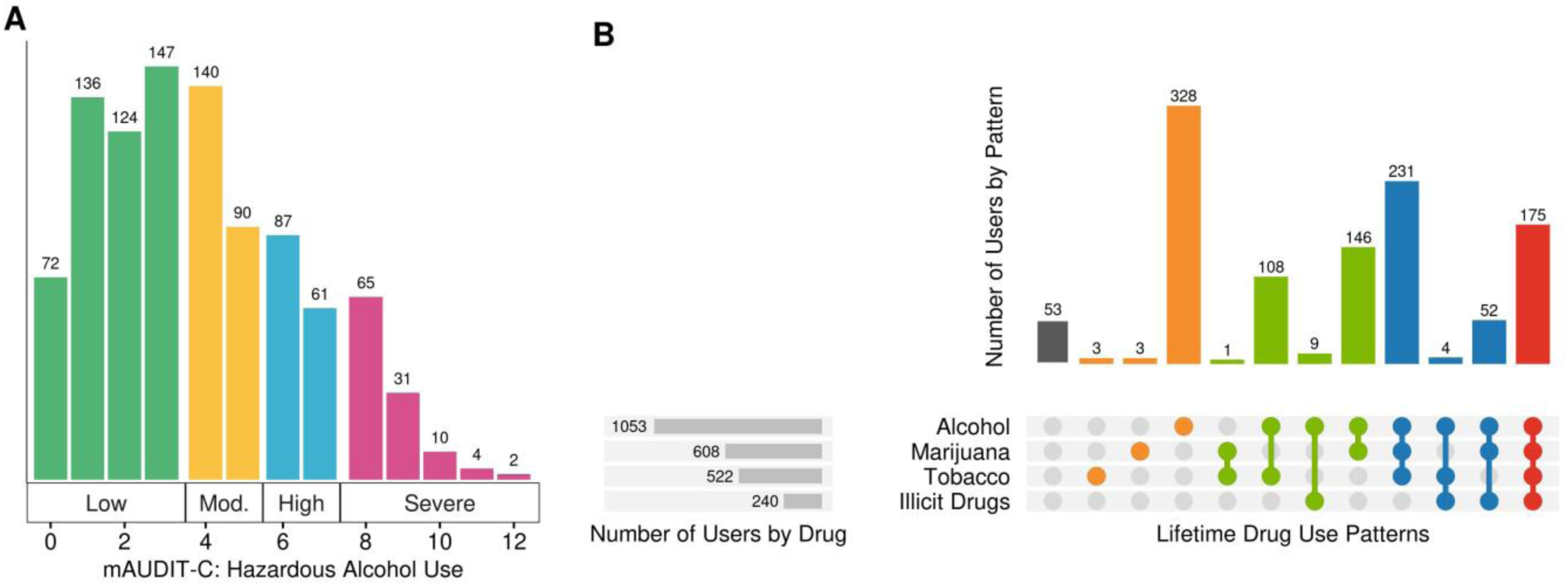
Substance Use Endorsement. A) Composite mAUDIT-C scores ranged from zero to 12 and were categorized into four levels of past-year hazardous risk: low (green; 0 to 3), moderate (yellow; 4 to 5), high (blue; 6 to 7), and severe (pink; 8 to 12). B) *Color indicates the number of drugs used across the lifetime.* Participant endorsement of having ever used any of the four major substance types are pictured. 65% (n = 726) of the sample endorsed a pattern of polysubstance use throughout the lifetime (i.e., 2+ substances).

### Associations of Substance Use with Brain Structure

Mixed linear effects models tested associations between mAUDIT-C and 24 measures of brain structure, including global cortical thickness and surface area, thickness of 8 bilateral cortical regions, and volume of 3 bilateral subcortical regions. Both global measures and 8 regional thickness measures survived FDR correction (*p_FDR_*<0.05) for multiple comparisons, with evidence for negative associations with thickness and positive associations with cortical surface area. The strongest association was with mean global cortical thickness (*β* = −0.12, *p* < 0.001) (**Figure 2**). Analyses were repeated with global thickness as a covariate to probe whether regional associations reflected the global effect or were local associations; no region survived FDR correction for multiple comparisons (**eFigure 4; Supplemental Data**). *Post-hoc* analyses tested associations between all regional measures of brain thickness, surface area, and volume and all substance use measures. Of 123 significant associations that survived correction for each substance use variable and brain measure modality, 111 (90.2%) were accounted for by inclusion of cortical thickness as a covariate (**eResults; eFigure 5; Supplemental Data**). No region survived FDR-correction for all tests in these *post-hoc* analyses.

**Figure 2.**
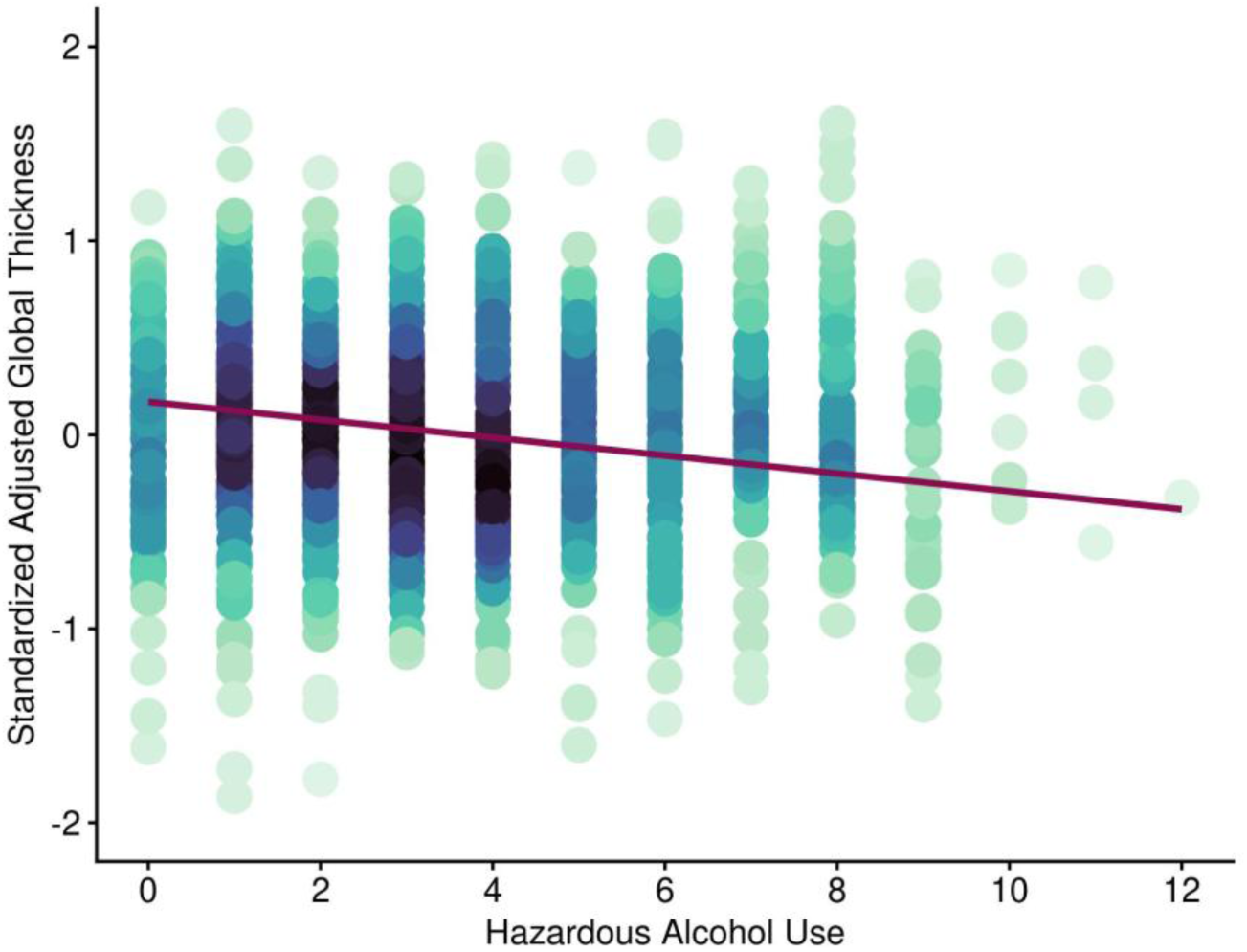
Hazardous Alcohol Use Predicts Attenuated Global Brain Thickness. Association of hazardous alcohol use (mAUDIT-C) with global cortical thickness, residualized for covariates and standardized (i.e., SD=1). Covariates included age, age^2^, sex, socioeconomic status, level of educational attainment, intracranial volume, and sibling status. Shading reflects the density of overlapping points. mAUDIT-C was uniquely associated with attenuated global brain thickness (*β* = −0.12, *p* < 0.001).

Analyses then tested the association of other primary substance use variables with global brain thickness. Significant (*p_FDR_*<0.05) associations included alcohol use age of onset (*β* = 0.08, *p* = 0.014), binary lifetime marijuana use (*β* = −0.12, *p* < 0.001), and binary lifetime tobacco use (*β* = −0.09, *p* = 0.001). Only mAUDIT-C and marijuana use remained significant when all four substance use variables were jointly entered into a single model (**Figure 3**; **Supplemental Data**). The addition of all other primary substance use variables did not improve fit over a model that included only mAUDIT-C and marijuana use (*p*=0.07). *Post-hoc* analyses tested a marijuana use and mAUDIT-C interaction, in addition to cortical thickness associations with secondary substance use variables (e.g., breathalyzer and urine drug screenings) and potential mediating variables (e.g., cognition, mental health, personality), finding no significant associations (**eResults**, **Figures e6 & e7; Supplemental Data**).

**Figure 3.**
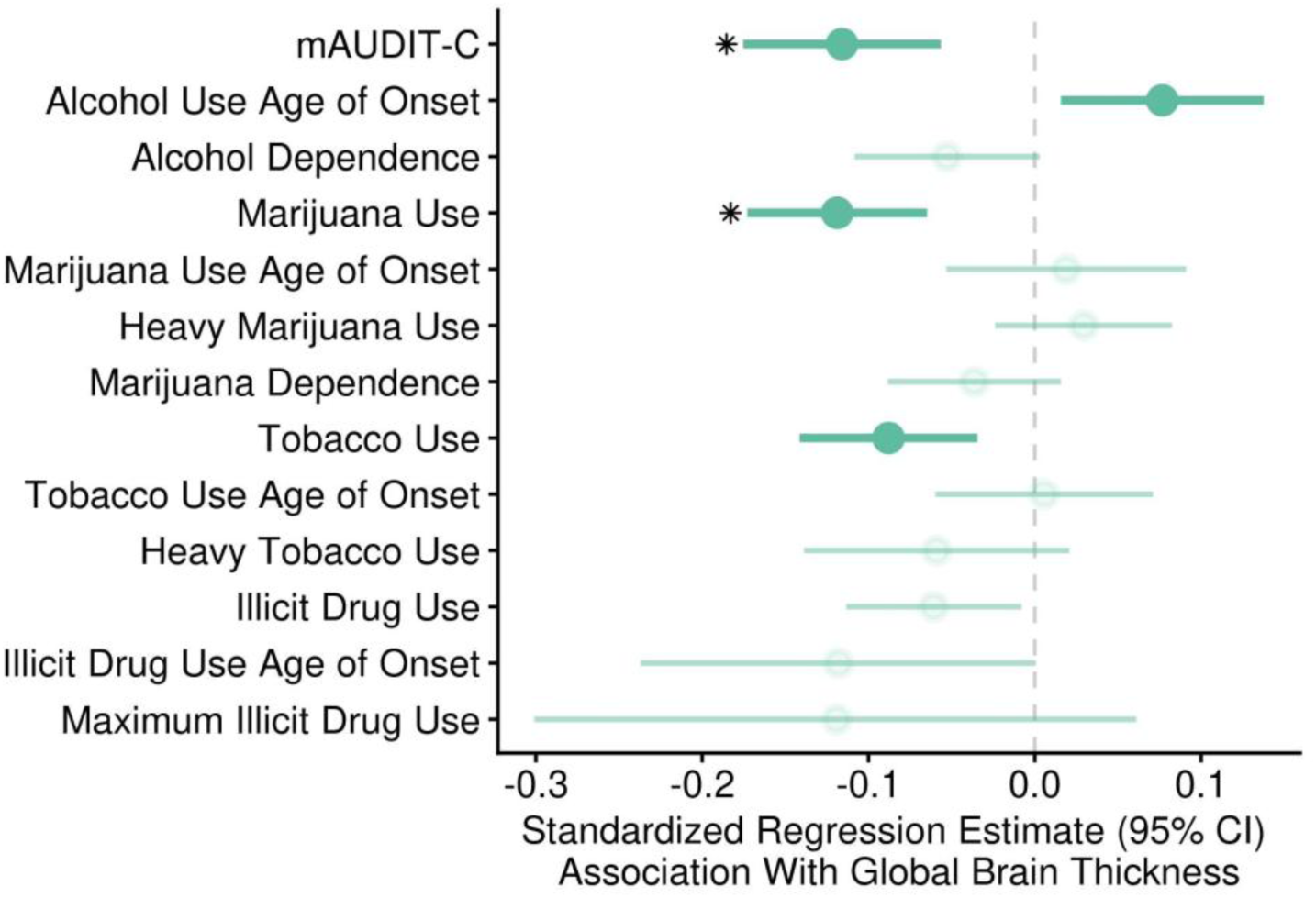
Shared- and Unique-Drug Associations with Attenuated Brain Thickness. Standardized regression estimates of alcohol, marijuana, tobacco, and illicit drug use variables predicting whole brain cortical thickness. Lines reflect the 95% confidence interval of the estimate. Note that these dimensions were not standardized across drug type, as collection of these data varied. *Bold indicates survival of multiple test correction and evidence for shared-effects on global brain thickness. Starred indicates drug-specific, unique effects on global brain thickness*.

### Probing Evidence for Predispositional and Exposure Effects

Within- and between-family analyses examined exposure and predispositional effects of substance use on brain structure. Analyses were repeated within increasingly stringent subsamples, i.e., full siblings only, twins only, and monozygotic twins only. Both mAUDIT-C (*β* = −0.11, *p* < 0.001) and marijuana use (*β* = −0.07, *p* = 0.002) evidenced a within-family effect on global brain thickness up to full siblings (**Figure 4A**). These associations remained significant after controlling for the alternative predictor (mAUDIT-C, *β* = −0.09, *p* < 0.001; marijuana use, *β* = −0.07, *p* = 0.004; **eFigure 8**), suggesting unique, within-family drug use effects on global brain thickness. Only marijuana use evidenced a between-family effect, which survived up to full siblings (*β* = −0.11, *p* = 0.007) and correction for mAUDIT-C (*β* = −0.09, *p* = 0.04; **Figure 4A; Supplemental Data**). These data survived FDR correction.

**Figure 4.**
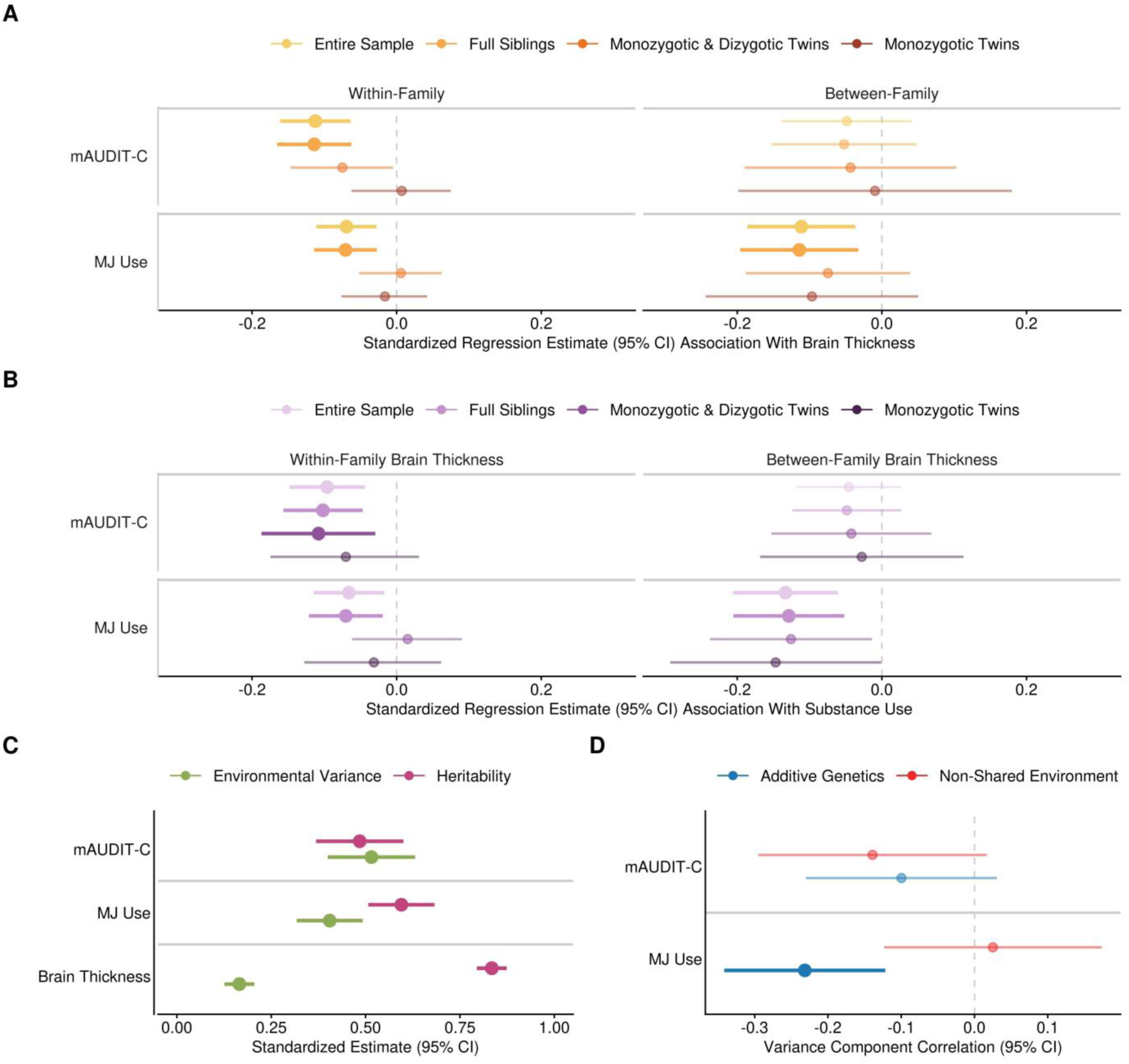
Drug Use Associations With Brain Structure Reflect a Combination of Predispositional Risk and Exposure Effects. A) Regression estimates of within- and between-family drug use variables fit to predict whole brain thickness are pictured. Y-axis variables are split by mAUDIT-C and Marijuana Use. X-axis data are split by within- (left) and between- (right) family estimates. Color denotes the sample included in the analysis. B) Regression estimates of within- and between-family whole brain thickness fit to predict substance use variables are pictured. Y-axis variables are split by mAUDIT-C and Marijuana Use, the two drugs evidencing unique-effects on whole brain thickness. X-axis data are split by within- (left) and between- (right) family whole brain estimates. Color denotes the sample included in the analysis. Bold indicates significance. C) Standardized regression estimates of environmental variance (green) and heritability (pink) fit to predict mAUDIT-C, Marijuana Use, and Brain Thickness are pictured. D) Variance component correlations of additive genetics (blue) and non-shared environment (red) between mAUDIT-C and Marijuana Use are pictured. Points reflect estimates, lines indicate 95% confidence intervals. *MJ Use = Marijuana Use*. *Bold indicates significance*.

As data are cross-sectional, within/between analyses are consistent with a bi-drectional relationship between substance use and brain structure. To further probe this interpretation, models tested the association of within- and between-family brain thickness with substance use. A within-family brain thickness effect was observed on mAUDIT-C (*β* = −0.11, *p* = 0.008) up to twins only and marijuana use (*β* = −0.07, *p* = 0.007) up to full siblings (**Figure 4B**). A between-family effect of brain thickness was observed on marijuana use (*β* = −0.13, *p* = 0.001) up to full siblings. These associations survived FDR correction (*p_FDR_*<0.05) (**Supplemental Data**).

SOLAR-Eclipse was used to decompose the correlation between cortical thickness and substance use into additive genetics (i.e., genetic correlation) and non-shared environment components. All variables were significantly heritable (mAUDIT-C: *h^2^r* = 0.48, *p* < 0.001; marijuana use: *h^2^r* = 0.59, *p* < 0.001; cortical thickness: *h^2^r* = 0.83, *p* < 0.001; **Figure 4C**). A significant genetic correlation between lifetime marijuana use and cortical thickness was observed (*ρG* = −0.23*, p* < 0.001; **Figure 4D**). This association survived correction for mAUDIT-C (*ρG* = −0.2*, p* = 0.004). No non-shared environmental correlations were observed.

## Discussion

Polysubstance use is a common pattern of drug use associated with exacerbated physical and mental health symptoms relative to monosubstance use ^5,6^. Despite this, the psychiatric neuroimaging literature has largely examined associations of brain structure within individual substance types. The current study investigated the shared- and unique-drug effects of alcohol, marijuana, tobacco, and illicit drug use on brain structure. In addition, family-based analyses probed whether these associations might be attributable to exposure or predispositional risk effects. Analyses found evidence for shared effects of alcohol, marijuana, and tobacco use on global cortical thickness, as well as unique effects of both alcohol and marijuana. These associations reflected a combination of substance exposure and predispositional risk effects. Overall, these results suggest that the negative consequences of polysubstance use may arise, in part, as a result of exposure to the additive effects of multiple substances.

As nearly the entire sample (95%) endorsed lifetime alcohol use, brain structures of interest were identified according to a previous review of alcohol-related associations influenced by exposure and/or predispositional risk effects ^24^. Analyses first examined hazardous alcohol use (mAUDIT-C) associations with these measures. Global brain thickness exhibited the largest association, and follow-up analyses found that it accounted for all other associations with mAUDIT-C (**eFigure 4**). This trend continued in *post-hoc* analyses where global brain thickness explained 90% of all possible substance use and brain structure associations (**eFigure 5**). In addition to hazardous alcohol use, analyses then found that lifetime marijuana use, lifetime tobacco use, and age of alcohol use onset were also associated with global brain thickness. These were shown to reflect, in-part, a shared-drug effect. These results are consistent with prior work identifying evidence for shared-drug effects across marijuana, tobacco, and alcohol use when examining measures of regional thickness ^15,16,19,46,47^. Further, they suggest that shared-drug effects are widespread across the brain.

In addition to a shared-drug effect, there were unique-drug effects of hazardous alcohol and lifetime marijuana use, such that they explained unique variance in global brain thickness that exceeded the other substance-use measures examined (**Figure 3**). The effects of these drugs were observed to be additive rather than multiplicative. While there is robust evidence for associations of alcohol and brain structure, we extend this literature with the identification of unique-effects in the context of multiple other substance types. The prior literature examining shared and unique effects of marijuana use is mixed. Our results align with findings of a unique-effect of lifetime marijuana use on regional brain thickness ^48–50^. In contrast, other investigations have reported non-significant thickness associations, shared-effects exclusively when examining chronic use, or that associations reflect within-person changes in use ^16,21,22,51^. The present results were not explained by broader polysubstance use or any other dimension of marijuana use, including the urine drug test for cannabis. The unique effect observed here may thus reflect an intermediate pattern of use (e.g., past six to twelve months), rather than recent, long-term, or chronic use, and is in need of further study. Overall, findings show that hazardous alcohol and lifetime marijuana use are associated with differences in brain thickness that exceed other drug types and dimensions of use.

To assess the influence of an unexplored third variable mediating the relationship between drug use and brain structure, exploratory analyses examined associations of all cognitive, mental, and physical health variables with global brain thickness, hazardous alcohol use, and marijuana use. Several nominally significant associations were found (**Figure e7**), but no effects consistent with mediation were observed (**eResults, Supplemental Data**). Additionally, sex differences were not observed.

We then sought to leverage the family-design of the Human Connectome Project to identify whether predispositional and/or exposure effects contribute to these unique associations. Findings support a bi-drectional model, in which substance use exhibits exposure effects on brain structure, and in which brain structure serves as a marker for risk factors which drive changes in substance use behavior. The observed within-family associations are consistent with the interpretation that hazardous alcohol and lifetime marijuana use each exert unique exposure effects on global cortical thickness ^48–50^. Equivalently, these are also consistent with a model in which cortical thickness drives, or serves as a marker for factors which drive changes in substance use behavior ^8,52^. These findings align with reports that brain structure recovers with substance abstinence ^53,54^, though further work is needed to determine guidance on the length of abstinence necessary for structural recovery.

An emerging literature has identified genetic and environmental risk factors that are associated with substance use and brain structure broadly ^9,10,14,16,24,49–52^. Consistent with this, associations between brain thickness and lifetime marijuana use observed here partially reflect predispositional risk factors, including genetic and shared-environmental risk factors, which were not explained by concurrent alcohol use. While prior work has reported non-significant genetic correlations between lifetime marijuana use and brain thickness, it is possible that differences in phenotyping or power (i.e., relatively few genome-wide significant loci) obscured this association ^14,50^. Additionally, previous investigations report predispositional risk effects contributing to the association of alcohol use with brain structure ^8,14^, including in a study of regional volume in the HCP ^28^. In contrast, a between-family effect and a genetic correlation between alcohol use and global cortical thickness was not observed in the present analyses. This discrepancy may reflect, in part, that imaging modalities differ in heritability ^55^, with evidence that cortical surface area and volume are more heritable than cortical thickness. Prior genetic analyses have also used data from large genome-wide association studies which, out of necessity, combine neuroimaging data across a wide range of ages (i.e., ages 3-90 ^55^). The relatively narrow age range of the present sample (ages 22-37) may attenuate or mask the expression of genetic risk effects that become apparent at younger, or older, ages.

### Limitations

The current study is characterized by some limitations. Analyses were restricted by the design of the parent study. The HCP aimed to evaluate cross-sectional individual differences in brain circuits, behavior, and genetics. As substance use was not a principal focus, data regarding dimensions of substance use were not standardized across drug types, and data on the quantity of substance used was largely unavailable. While the HCP is a large adult twin neuroimaging study, we did not have the power to disentangle shared environment and genetic effects in the present analyses. Most participants were White (*n* = 862, 74.7%), despite only making up 44% of the surrounding locale where data were collected ^56^. Further research should aim to address these gaps to provide stronger evidence for causal and/or predispositional effects, contribute to standardized operationalization, and increase generalizability.

## Conclusion

Shared- and unique-drug effects of alcohol and marijuana are associated with attenuated global brain thickness. These bidirectional associations reflect a mix of causal and predispositional effects, consistent with both neurobiological and neurodevelopmental models of addiction ^24^. Future work combining twin modeling with longitudinal designs and representative samples will aid in understanding the underlying mechanisms of how substance use influences the brain.

## Acknowledgements

This study was supported by R00AA030808 (DAAB). Data were provided by the Human Connectome Project, WU-Minn Consortium (Principal Investigators: David Van Essen and Kamil Ugurbil; 1U54MH091657) funded by the 16 NIH Institutes and Centers that support the NIH Blueprint for Neuroscience Research; and by the McDonnell Center for Systems Neuroscience at Washington University. This research was completed in part with computational resources and technical support provided by the Research Computing Center at the Medical College of Wisconsin.

## Data availability

Data used in the preparation of this article were obtained from the Human Connectome Project (HCP) Young Adult, which includes imaging data on over 1100 healthy young adults, available at https://www.humanconnectome.org/.

## Code availability

Analysis code is available at https://github.com/BEARLabScience/BrainStructureSubstanceUse

