## Supplement for "Brain Structure and Substance Use: Disentangling Risk, Exposure, and Drug-Specific Effects"

**Table of Contents:**


| | Page |
|---|---|
| eMethods | 3 |
| Measures | 3 |
| Hazardous Alcohol Consumption | 3 |
| Brain Structure | 3 |
| eData Analytic Plan | 3 |
| Mediation and Moderation Analyses | 3 |
| Whole-brain Analyses | 3 |
| eResults | 5 |
| Exploratory Mediation Analyses | 5 |
| Exploratory Moderation Analyses | 5 |
| Exploratory Whole Brain Analyses | 5 |
| Supplemental References | 7 |
| eTable 1. SSAGA to mAUDIT-C Transformation. | 8 |
| eTable 2. Substance Use Endorsement Across Dimensions. | 9 |
| eFigure 1. Sample Characteristics. | 10 |
| eFigure 2. Patterns of Hazardous Use Across Drug Types. | 11 |
| eFigure 3. Drug Use Frequency Throughout the Lifetime. | 12 |
| eFigure 4. Global Brain Thickness Explains Regional Alcohol Effects on Brain Structure. | 13 |
| eFigure 5. Cortical Thickness Accounts for the Majority of Substance Use Associations. | 14 |
| eFigure 6. Secondary Drug Use Variable Associations with Whole Brain Thickness. | 15 |
| eFigure 7. Exploratory Analyses of Possible Mediators. | 16 |
| eFigure 8. Unique Within- and Between-Family Drug Use Effects on Global Brain Thickness. | 17 |

**eMethods**

Young adult siblings, primarily twins, were recruited from the local community by staff at Washington University in St. Louis, Missouri. Ineligibility was defined as having a significant history of neurological, psychiatric, or cardiovascular disease. For further details on inclusion and exclusion criteria, see Van Essen et al., 2013. Study Day 1 involved signature of the informed consent and a structural MRI. Cognitive, physical, and self-report assessments were completed. Study Day 2 consisted of biological drug tests and compensation. Finally, a phone interview regarding the history of substance use was conducted. Each participant received $400 remuneration, as well as additional winnings ($5) and travel expenses. Data were accessed by the current research team for analysis, as approved by the Medical College of Wisconsin IRB.

**Measures.**

***Hazardous Alcohol Consumption***

As in prior work [1], responses to three items in the SSAGA that are similar to questions in the AUDIT-C were scored and summed to generate a modified AUDIT-C (mAUDITC). These items were: 1) *On how many days did you drink any beverages containing alcohol during the last 12 months?*; 2) *On a typical day, how many drinks did you consume on average?*; and, 3) *Now I'd like you to think about the last 12 months. How often did you have 5 or more drinks in a 24-hour period?* See **eTable 1** for scoring details.

***Brain Structure***

The acquisition and processing details of this dataset are described in detail in prior reports. Briefly, relevant steps for this study from the HCP processing pipeline within FreeSurfer v5.3.0 included: 1) spline-based down-sampling of the 0.7mm T1 image to 1mm; 2) intensity normalization and Talairach transformation; 3) skull registration; 4) skull stripping; 5) subcortical segmentation; 6) creation of white and pial surfaces and their refinement using the full (0.7mm) resolution data; 7) refinement of the pial surface using the T2-SPACE scan to help exclude cerebral spinal fluid (CSF) and dura, 8) extraction of cortical thickness and surface areas estimates from the Desikan atlas [2] parcellation, and 9) extraction of subcortical volume estimates from the FreeSurfer ASEG atlas [3–5].

**eData Analytic Plan.**

***Mediation and Moderation Analyses***

Exploratory analyses aimed to identify mediating variables that could explain the relation between unique substance use variables and brain structure. All behavioral, cognitive, and mental health variables not previously analyzed were fit to predict brain ROIs and substance use variables evidencing unique effects. These variables, unrelated to quantifying substance use (e.g., working memory tasks, sleep quality indexes, visual acuity) were then fit to mediate the relation between substance use and brain structure.

To test multiplicative effects on brain structure, interactions between substance use variables were fit to predict brain ROI. Then, as a test of potential sex differences, interactions between gender and unique substance use variables were fit to predict brain ROI. Finally, age and birth control were fit to interact and predict brain ROI as an investigation of the influence of a commonly used contraceptive.

***Whole-brain Analyses***

The preliminary post-hoc analysis began with examining the associations between biological drug tests (i.e., marijuana, methamphetamine, cocaine, breathalyzer, oxycontin, opiates, and amphetamine) and global brain thickness via mixed effects models. The number of times marijuana and tobacco were used was also examined. As the distribution of these data were irregular (i.e., heavily zero-inflated or bimodal), they were not included as primary variables of interest. These analyses were corrected using the FDR method.

The final post-hoc analysis included three sets of mixed linear effects models to evaluate the accuracy of selected ROI. First, all drug variables were fit to predict all measures of brain structure included in the HCP, with the same covariates listed in the **Data Analytic Plan**. Second, the previous set of analyses were repeated with global brain thickness as an additional covariate. Finally, all drug variables were fit to predict only the ROI included in the primary analyses (**eFigure 4**). These sets of analyses included three levels of FDR correction. The first level adjusted within each modality and drug type combination (e.g., volume by marijuana separate from volume by tobacco). The second level adjusted per drug type (e.g., all analyses with marijuana dependence jointly corrected). The final level adjusted across all analyses at once.

**eResults**

**Exploratory Mediation Analyses.**
Mixed linear effects models explored potential variables that might mediate the association between global thickness and substance use. First, models tested the association of all available cognitive, emotional, and behavioral variables with global brain thickness. Of the 275 variables tested, 17 were nominally significant, but none survived multiple test correction: all working memory trials accuracy ($\beta$ = -0.06, $p$ = 0.021), median reaction time across working memory trials ($\beta$ = 0.05, $p$ = 0.045), non-target working memory trials accuracy ($\beta$ = -0.06, $p$ = 0.022), target working memory trials accuracy ($\beta$ = -0.05, $p$ = 0.046), average difficulty level across language and math tasks ($\beta$ = -0.05, $p$ = 0.048), oral reading recognition ($\beta$ = -0.07, $p$ = 0.021), NIH crystallized cognition ($\beta$ = -0.08, $p$ = 0.024), relational task accuracy ($\beta$ = -0.06, $p$ = 0.042), Penn Line Spatial Orientation median reaction time ($\beta$ = 0.06, $p$ = 0.022), neutral emotion recognition ($\beta$ = 0.07, $p$ = 0.010), NIH sadness ($\beta$ = 0.06, $p$ = 0.014), perceived hostility ($\beta$ = 0.08, $p$ = 0.002), PSQI other sleep problems ($\beta$ = 0.09, $p$ < 0.001), SSAGA childhood conduct issues ($\beta$ = 0.06, $p$ = 0.031), ASR rule breaking ($\beta$ = -0.06, $p$ = 0.043), visual acuity ($\beta$ = 0.09, $p$ < 0.001), and birth control use ($\beta$ = -0.09, $p$ = 0.017) [**Supplemental Data**]. See **eFigure 4** for visualization of these associations.

Next, models tested whether these variables were associated with mAUDIT-C and marijuana use. Significant predictors of the former include average difficulty level across language and math tasks ($\beta$ = 0.09, $p$ = 0.004), SSAGA childhood conduct issues ($\beta$ = 0.08, $p$ = 0.011), ASR rule breaking ($\beta$ = 0.32, $p$ < 0.001), visual acuity ($\beta$ = -0.07, $p$ = 0.012), and birth control use ($\beta$ = 0.10, $p$ = 0.040). Significant predictors of the latter include SSAGA childhood conduct issues ($\beta$ = 0.08, $p$ = 0.011), ASR rule breaking ($\beta$ = 0.30, $p$ < 0.001), and birth control use ($\beta$ = 0.12, $p$ = 0.005). As ASR rule breaking showed the largest and most consistent associations, mediation models [6] formally tested the mediation, which was not significant for both alcohol and marijuana ($\beta$ = -0.06, $p$ = 0.4) [**Supplemental Data**].

**Exploratory Moderation Analyses.**
An interaction between mAUDIT-C and Marijuana Use was tested as a predictor of global brain thickness. This was non-significant ($\beta$ = 0.03, $p$ = 0.6). Additionally, the interaction of gender with substance use, predicting thickness, was explored. Both interactions of gender with mAUDIT-C ($\beta$ = -0.05, $p$ = 0.1) and Marijuana Use ($\beta$ = -0.005, $p$ = 0.8) were non-significant.

**Exploratory Whole-brain Analyses.**
Analyses examined associations of drug tests (i.e., marijuana, methamphetamine, cocaine, breathalyzer, oxycontin, opiates, and amphetamine), as well as the number of times marijuana and tobacco were used, with global brain thickness via mixed effects models. None of these associations were significant. Data are visualized in **eFigure 3**.

Mixed linear effects models were fit to test the association between all drug variables and all brain structures included in the data set. 123 associations survived the first level of FDR correction detailed in eData Analytic Plan, ranging across nine drug use variables and 63 brain structure measures (**eFigure 5**). Ten survived the second level: mAUDIT-C with choroid plexus volume ($\beta$ = 0.13, $p$ < 0.001); number of times used illicit drugs with isthmus cingulate area ($\beta$ = -0.33, $p$ < 0.001); marijauna user with predicted global brain thickness ($\beta$ = -0.12, $p$ < 0.001), as well as regional thickness of the left caudal middle frontal lobe ($\beta$ = -0.12, $p$ < 0.001), left ($\beta$ = -0.12, $p$ < 0.001) and right ($\beta$ = -0.11, $p$ < 0.001) fusiforum gyrus, left inferior temporal lobe ($\beta$ = -0.11, $p$ < 0.001), left superior temporal lobe ($\beta$ = -0.11, $p$ < 0.001), left supramarginal gyrus ($\beta$ = -0.11, $p$ < 0.001), and right lateral occipital lobe ($\beta$ = -0.10, $p$ < 0.001). Only the five strongest associations survived the third level of FDR correction.

The previous analyses were repeated with global brain thickness as an additional covariate. Notably, global brain thickess and intracranial volume were weakly correlated ($r = 0.2$). The following associations remained significant after the first level of FDR correction: number of times used illicit drugs with the left isthmus cingulate area ($\beta = -0.31$, $p < 0.001$) and thickness ($\beta = 0.30$, $p < 0.001$); lifetime illicit drug use with right superior frontal ($\beta = -0.06$, $p = 0.001$), left transverse temporal ($\beta = 0.09$, $p = 0.001$), and left caudal middle frontal ($\beta = -0.06$, $p = 0.002$) lobe thicknesses; the methamphetamine drug test predicted right inferior parietal ($\beta = -0.07$, $p < 0.001$) and left pericalcarine ($\beta = 0.08$, $p = 0.001$) area; a breathalyzer test of over 0.08 with e right cerebellum cortex volume ($\beta = 0.07$, $p < 0.001$) and left lateral orbitofrontal thickness ($\beta = 0.08$, $p < 0.001$); mAUDIT-C with left choroid plexus volume ($\beta = 0.12$, $p < 0.001$); the age of onset of alcohol use with left caudate volume ($\beta = -0.09$, $p < 0.001$); finally, lifetime tobacco use with volume of the right amygdala ($\beta = -0.07$, $p < 0.001$). Only the strongest association survived the second and third level of correction.

Finally, analyses examined the association of all drug variables with only the brain ROIs originally selected for analysis (i.e., those identified in a prior review), with global brain thickness as a covariate. Nine associations remained significant after accounting for the first level of FDR correction: binary lifetime tobacco use with right amygdala ($\beta = -0.07$, $p < 0.001$), left hippocampus ($\beta = -0.06$, $p = 0.008$), and right hippocampus ($\beta = -0.06$, $p = 0.004$) volume; binary lifetime illicit drug use with the right superior frontal ($\beta = -0.06$, $p = 0.001$) and left caudal middle frontal ($\beta = -0.06$, $p = 0.002$) thickness; the biological cocaine drug test with the left hippocampus volume ($\beta = 0.06$, $p = 0.003$); breathalyzer test with the volume of the left ($\beta = 0.06$, $p = 0.008$) and right ($\beta = 0.07$, $p < 0.001$) cerebellum cortex; finally, number of times used marijuana with left superior frontal thickness ($\beta = -0.09$, $p = 0.002$). Only the first association survived the second level of correction. None of the associations survived the third level of correction (i.e., correction for all tests). These three tests are visualized in **eFigure 2**.

| **AUDIT-C** | **SSAGA** |
|---|---|
| 1. How often did you have a drink containing alcohol in the past year?<br>a) Never (0 pt)<br>b) Monthly or less (1 pt)<br>c) 2 to 4 times per month (2 pts)<br>d) 2 to 3 times per week (3 pts)<br>e) 4 or more times per week (4 pts) | 1. On how many days did you drink any beverages containing alcohol during the last 12 months?<br>a) Never (0 pt)<br>b) 1 to 2 days per year (1 pt)<br>c) 3 to 5 days per year (1 pt)<br>d) 1 day per month (1 pt)<br>e) 2 days per month (2 pts)<br>f) 3 days per month (2 pts)<br>g) 1 day per week (2 pts)<br>h) 2 days per week (3 pts)<br>i) 3 days per week (3 pts)<br>j) 4 days per week (4 pts)<br>k) 5 to 6 days per week (4)<br>l) Every day (4 pts) |
| 2. On days in the past year when you drank alcohol, how many drinks did you typically drink?<br>a) 0, 1, or 2 (0 pt)<br>b) 3 or 4 (1 pt)<br>c) 5 or 6 (2 pts)<br>d) 7-9 (3 pts)<br>e) 10 or more (4 pts) | 2. On a typical day, how many drinks did you consume on average?<br>a) 0, 1, or 2 (0 pt)<br>b) 3 or 4 (1 pt)<br>c) 5 or 6 (2 pts)<br>d) 7-9 (3 pts)<br>e) 10 or more (4 pts) |
| 3. How often did you have 6 or more (for men) or 4 or more (for women and everyone 65 and older) drinks on an occasion in the past year?<br>a) Never (0 pt)<br>b) Less than monthly (1 pt)<br>c) Monthly (2 pts)<br>d) Weekly (3 pts)<br>e) Daily or almost daily (4 pts) | 3. Now I'd like you to think about the last 12 months. How often did you have 5 or more drinks in a 24-hour period?<br>a) Never (0 pt)<br>b) 1 to 2 days per year (1 pt)<br>c) 3 to 5 days per year (1 pt)<br>d) 6 to 11 days per year (1 pt)<br>e) 1 day per month (2 pts)<br>f) 2 days per month (2 pts)<br>g) 3 days per month (2 pts)<br>h) 1 day per week (3 pts)<br>i) 2 days per week (3 pts)<br>j) 3 days per week (3 pts)<br>k) 4 days per week (3 pts)<br>l) 5 to 6 days per week (4 pts)<br>m) Every day (4 pts) |
| Sum = AUDIT-C | Sum = mAUDIT-C |

**eTable 1. SSAGA to mAUDIT-C Transformation.**
To capture hazardous alcohol use, items from the SSAGA were transformed to closely resemble those of the AUDIT-C, a well-validated and commonly used index of alcohol use. The items and response options of the AUDIT-C (left) and SSAGA (right) are listed. The original scoring of the AUDIT-C and adapted scoring of the SSAGA are denoted next to each item in parentheses.

| | *Range* | *M* (*SD*) |
|---|---|---|
| Age of Drug Use Onset | | |
| Alcohol | 5-31 | 17.69(3) |
| Marijuana | 7-34 | 17.98(3.5) |
| Tobacco | 5-33 | 16.38(3.8) |
| Illicit Drugs | 13-35 | 19.8(4.1) |
| Maximum Use | | |
| Illicit Drugs | 1-31 | 10.3(10.3) |
| | ***n(%)*** | |
| DSM-5 Dependency | | |
| Alcohol | 63(7) | |
| Marijuana | 102(9) | |

**eTable 2. Substance Use Endorsement Across Dimensions.**
Age of drug use onset is measured in years for all substances. Maximum illicit drug use is defined as the longest period of almost every day use and measured in days. Information related to DSM-5 Dependency are presented only for those who met criteria.

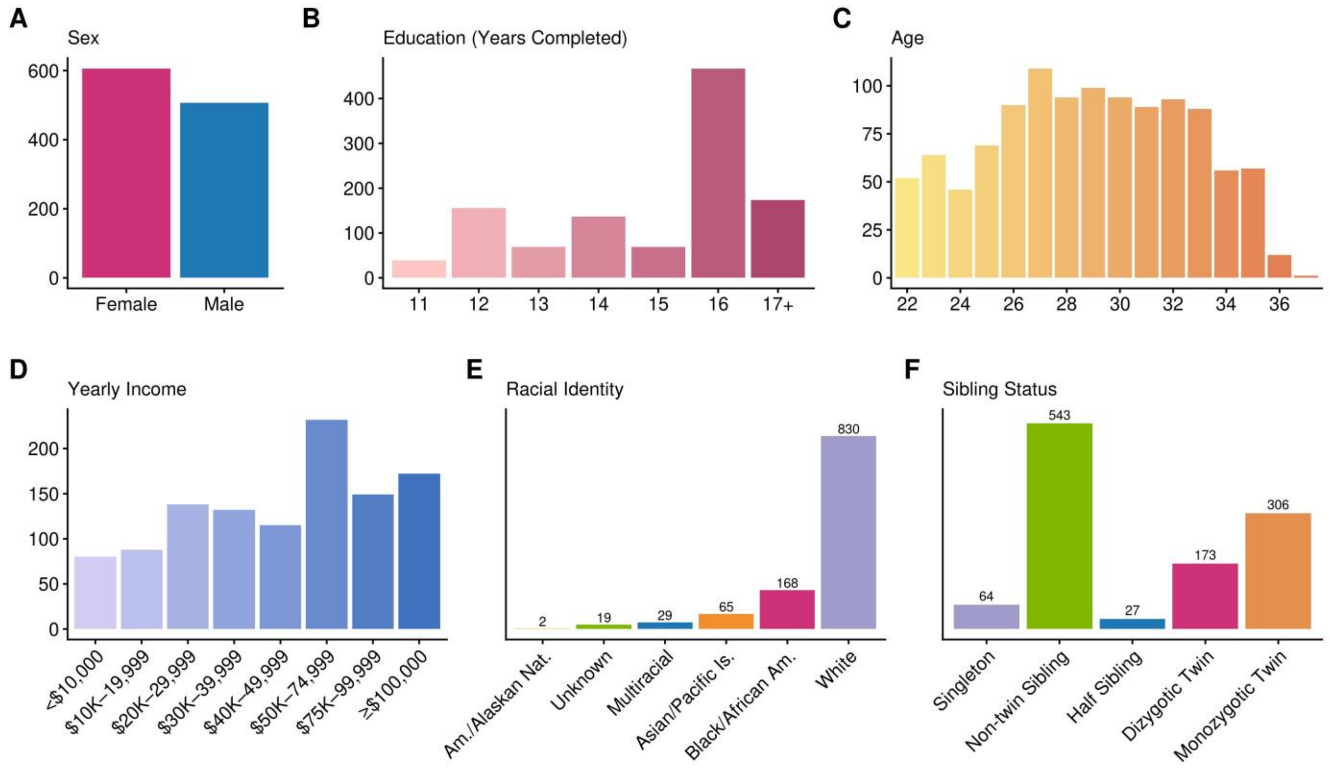


**eFigure 1. Sample Characteristics.**
Each plot visualizes a characteristic of the sample: A) self-reported sex, B) education measured in years completed, C) age in years, D) yearly income, E) self-reported racial identity, and F) sibling status as denoted via genetic testing and self-report.

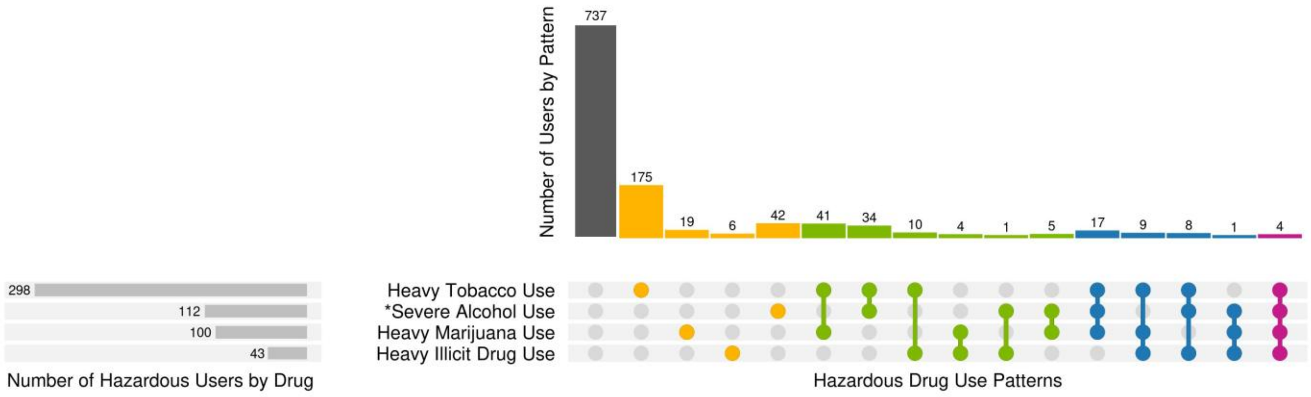


**eFigure 2. Patterns of Hazardous Use Across Drug Types.**
**indicates past-year measurement. All other variables are measured across the lifetime. Color denotes the number of drugs used hazardously.* Heavy marijuana and tobacco users were defined as having used marijuana more than 1,000 times and smoking more than 100 cigarettes, respectively. Heavy illicit drug use indicates at least four days of consecutive illicit drug use throughout the lifetime. Individuals with an mAUDIT-C score between 8 and 12 were categorized as severe alcohol users.

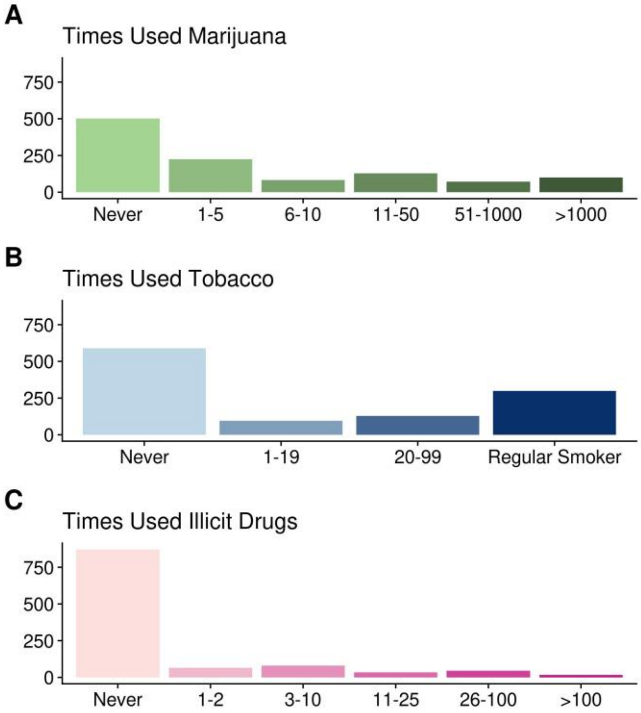


**eFigure 3. Drug Use Frequency Throughout the Lifetime.**
A), B), and C) denote the cumulative number of times used marijuana, tobacco, and illicit drugs, respectively. These data were collected in the Semi-Structured Assessment for the Genetics of Alcoholism (SSAGA) [7], capturing information on their history of alcohol, marijuana, tobacco, and illicit drug (i.e., cocaine, stimulant, sedative, opiates, other) use.

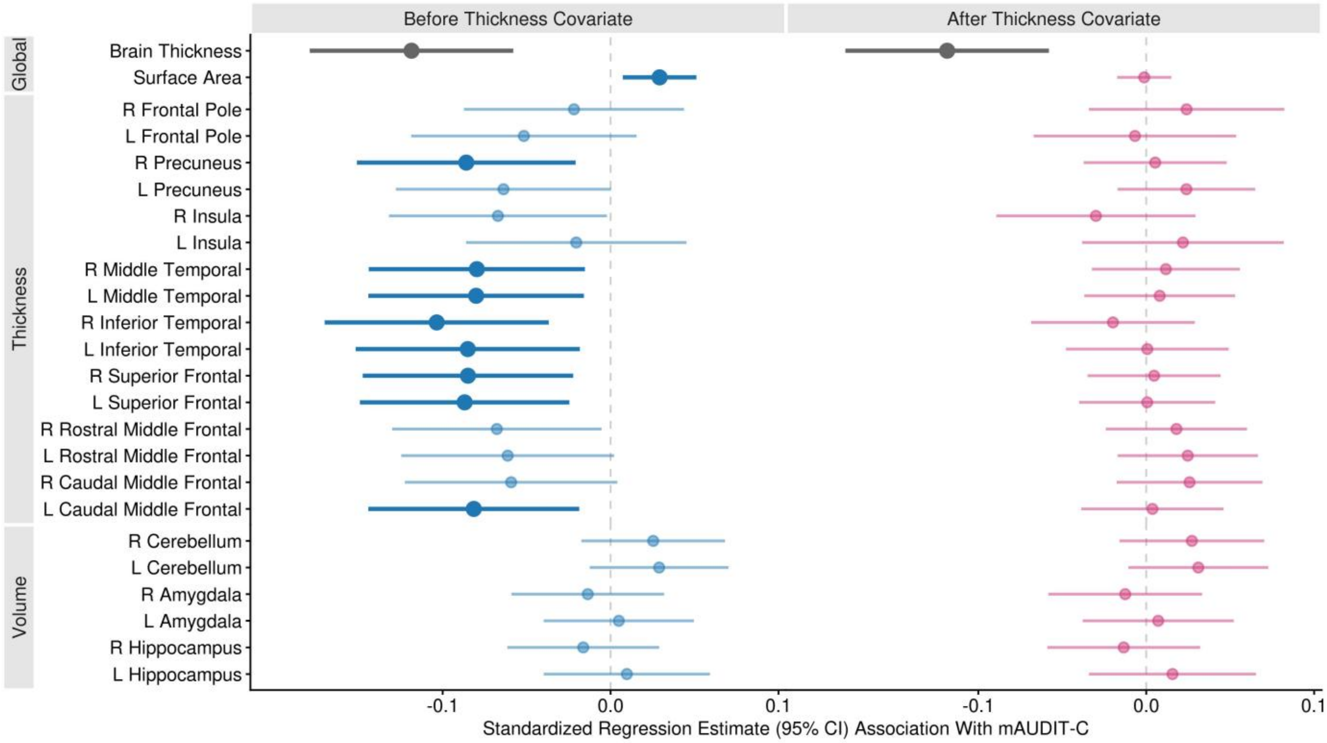


**eFigure 4. Global Brain Thickness Explains Regional Alcohol Effects on Brain Structure.** Standardized regression estimate associations between brain ROI and mAUDIT-C are pictured. Brain structures on the y-axis are split by modality, i.e., volume, thickness, and global measures. X-axis panels are split before (left, blue) and after (right, red) controlling for global brain thickness (grey). *Bold indicates significance following multiple test correction.*

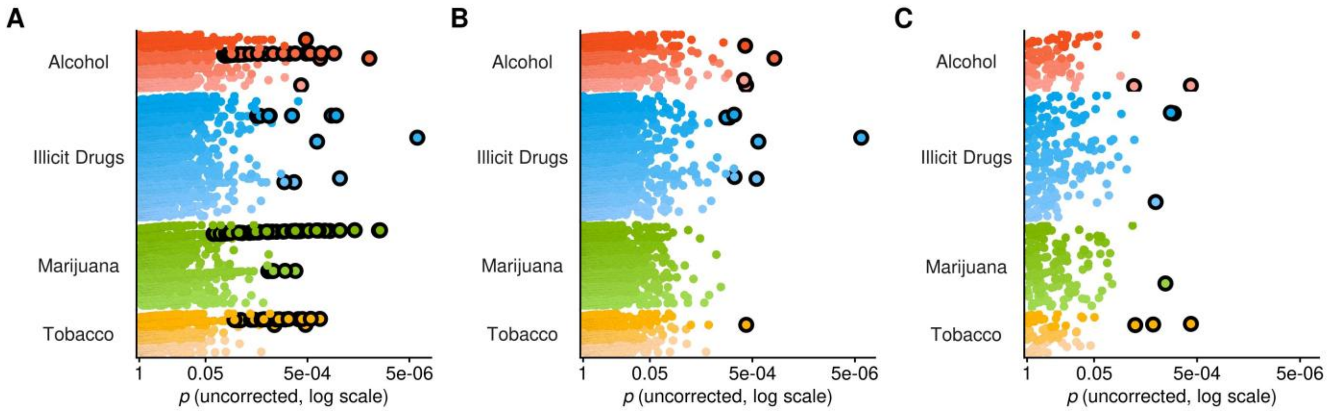


**eFigure 5. Cortical Thickness Accounts for the Majority of Substance Use Associations.**
A) The *p* values of associations between all available drug variables and brain measures before controlling for global brain thickness are pictured. Most of the significant drug variable associations are also strongly associated with global brain thickness, including mAUDIT-C and lifetime marijuana use. B) Analyses pictured in Panel A were repeated with global cortical thickness as an additional covariate. C) The *p* values of associations between the primary brain measures (i.e., those identified in a prior review) and all substance use variables of interest are pictured. These associations accounted for global brain thickness. *Colors indicate drug type. Shades of each color indicate a dimension of use (e.g., dependency, lifetime use, drug test). Individual points represent the p value of the association between a drug variable and a brain measure. Points outlined in black indicate survival of multiple test correction*.

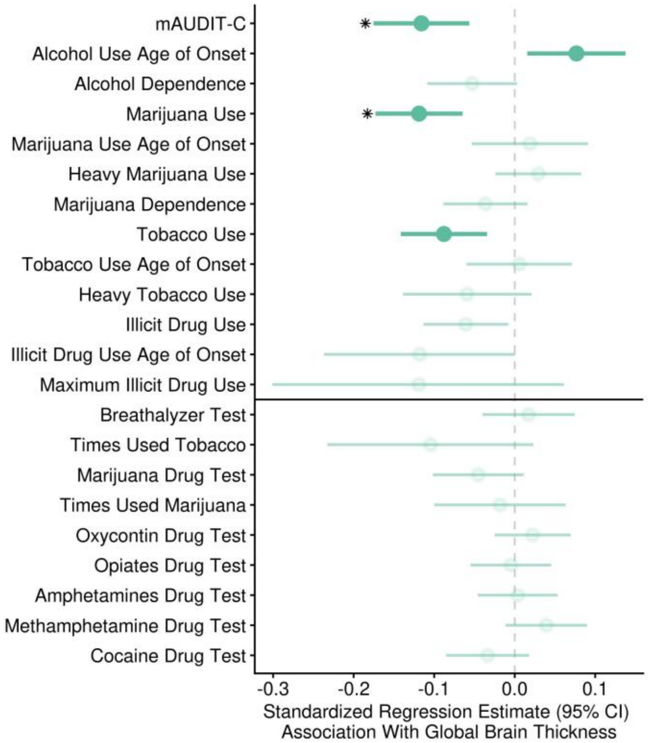


**eFigure 6. Secondary Drug Use Variable Associations with Whole Brain Thickness.** Participants completed biological drug screening to capture recent history of cocaine, marijuana, opiate, amphetamine, methamphetamine, and/or oxycontin use. Additionally, a breathalyzer test was administered. Scores above .08 were recorded. These associations with whole brain thickness, in addition to times used tobacco and marijuana, were calculated. Marijuana Use survives correction for the marijuana drug test; the two variables were moderately correlated (*r* = 0.3). ** indicates unique-drug effects. Bolded indicates significant shared-drug effects.*

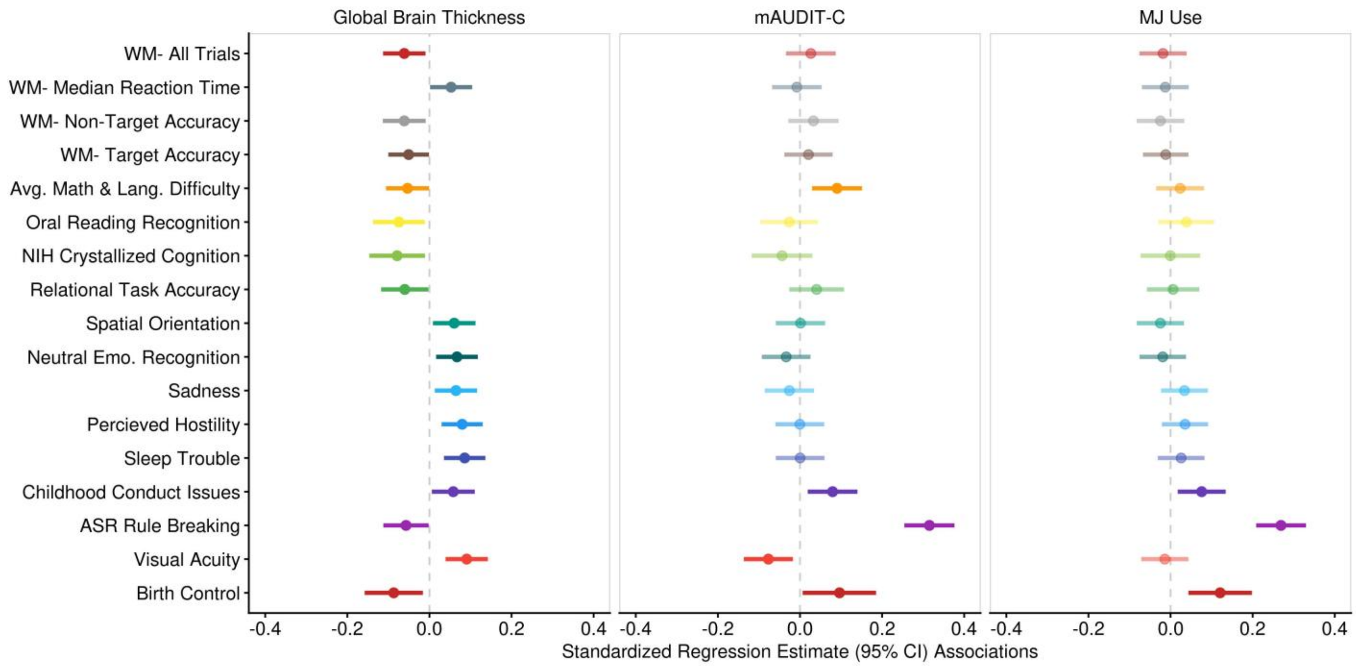


**eFigure 7. Exploratory Analyses of Possible Mediators.**
Significant predictors of brain thickness, mAUDIT-C, and Marijuana Use are pictured. None of the associations survived multiple test correction or were observed to have mediating effects on the relationship between global brain thickness and substance use. *Bold indicates significance.*

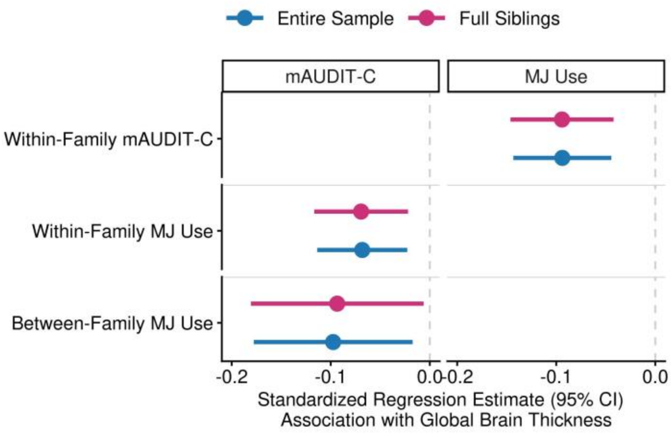


**eFigure 8. Unique Within- and Between-Family Drug Use Effects on Global Brain Thickness.** Marijuana use and mAUDIT-C evidenced within-family effects on cortical thickness up to a sub-sample of full siblings only. Unique effects of these associations were tested by controlling for the alternative drug predictors, i.e., mAUDIT-C (left) and marijana use (right). *Blue indicates that the entire sample was included in the model, while red denotes a sub-sample of only full siblings.*
